# Implementation of early child development and maternal mental health care package for forcibly displaced Myanmar population in Bangladesh refugee camps: a process evaluation and cost-effectiveness study

**DOI:** 10.64898/2026.10.01.26364288

**Authors:** Nida Khan, Muhammad Amir Khan, Khaleda Islam, Muhammad Ahmar Khan, Shaheer Ellahi Khan, Zunayed Al Azdi, Ishrat Jahan, Rumana Huque

**Affiliations:** Association for Social Development, Islamabad, Pakistan; ARK Foundation, Bangladesh; Health Services Academy, Islamabad, Pakistan

**Author notes:** **Corresponding author:** Email ID.

**Keywords:** Early child development, mental health, contextualised healthcare intervention, refugee camp, community-based providers

## Abstract

**Background:** The risk of developmental delays among infants in the refugee camps is high due to prolonged exposure to adverse situations such as poverty, violence, lack of access to basic necessities, safety, poor maternal mental health and inadequate access to healthcare. To reduce the risk of developmental delays, a contextualized early child development and maternal mental health care package, was implemented through enabling and engaging the refugee women as community-based care providers.

**Methods:** The process evaluation study was conducted using a mix-method design to explore the acceptability, feasibility, appropriateness, adoption, and cost of the Early Child Development (ECD) and maternal mental health care package in refugee camps, by applying Proctor et al’s framework. Data on quantitative indicators was reviewed for 348 mother-child dyads in the intervention arm. Qualitative interviews were conducted with 12 mothers, 5 community health workers, and 2 members of the implementing research team. Incremental Cost-Effectiveness Ratio (ICER) was calculated using the healthcare provider perspective to estimate the incremental cost-effectiveness of the ECD care package in reducing proportion of children with two or more developmental delays.

**Results:** Home-based delivery of counselling sessions was a key facilitator in ensuring the successful implementation of intervention. The delivery of intervention by modestly educated refugee women reduced cultural and language barriers and facilitated the home-based delivery of sessions. Mothers found the recommended activities feasible to do in routine as they were culturally appropriate, and did not require maternal literacy or additional resources. The intervention was cost-effective with an ICER of 63 USD in reducing the proportion of children with two or more developmental delays by one percentage point.

**Conclusion:** Contextualized ECD and maternal health care package is a potentially replicable and scalable intervention for refugee healthcare systems.

## 1. Introduction

The political conflict in Myanmar resulted in around 700,000 Myanmar nationals fleeing the country and seeking refuge in Bangladesh (1). Around 55% of these forcibly displaced Myanmar population in Bangladesh are children (2). Prolonged exposure to adverse situations such as poverty, violence, lack of access to basic necessities and poor maternal mental health places these children at high risk of developmental delays(3-5). Healthcare challenges such as shortage of healthcare resources in the refugee camps and cultural barriers between the service providers and the refugee population further exacerbate the risk of poor maternal mental health and developmental delays among children(6, 7). These challenges highlight the need to implement contextualized and potentially replicable model of early child development care, including maternal mental health, through enabling and engaging the refugee women. Integrated childhood development care interventions that are implemented within first 2 years of a child’s life have proven to be effective in preventing child developmental delays in low- and middle-income countries like Pakistan and Bangladesh(8-10).

To alleviate the burden of child developmental delays among forcibly displaced Myanmar nationals, we implemented a contextualized intervention package for early child development and maternal mental health in refugee camps of Bangladesh(11). This intervention was originally developed and evaluated in Pakistan(8) and was adapted according to the refugee population in consultation with an advisory group comprised of experts from Pakistan, Bangladesh and mothers of infants. The intervention was contextualized to suit the unique socio-cultural preferences, language, and lifestyle of refugee population and was delivered in community settings through recruiting women from the respective refugee camps of Bangladesh. A cluster randomised controlled trial has established the effectiveness of this integrated care package in preventing child development delays (publication in process). In line with Medical Research Council recommendations (12), a process evaluation study was conducted to get insights into the implementation process to explore what worked, how, and why.

Information on the acceptability, feasibility, adoption, appropriateness and cost-effectiveness of the intervention is to assist policymakers to make informed recommendations about the potential replicability and scale-up of the intervention in resource-constrained settings of Bangladesh.

The study has the following objectives:

1. To explore the acceptability, feasibility, appropriateness, adoption, and cost of the Early Child Development (ECD) and maternal mental health care package in refugee camps.
2. To estimate the cost-effectiveness of the ECD care package in preventing child developmental delays in refugee camps.

## 2. Materials and Methods

### 2.1. Study Design

The process evaluation and the cost-effectiveness were embedded within a parallel, two arms, cluster Randomized Controlled Trial (cRCT) (ISRCTN Registry ISRCTN10892553) (11). The process evaluation study was conducted using a mix-method design to explore the acceptability, feasibility, appropriateness, adoption, and cost of the ECD and maternal mental health care package in refugee camps.

### 2.2. Settings and Participants

The cRCT was conducted in refugee camps in Cox’s Bazar in Bangladesh. The study site was Kutupalong camp, with 20 camps, having definite boundaries and segments called ‘blocks.’ The number of blocks ranges from three to seven for a camp. Two blocks are combined to form a cluster. This study was conducted in the intervention arm (i.e., 11 clusters).

Data on quantitative indicators for the study was reviewed for 348 mother-child dyads in the intervention arm. We conducted qualitative interviews with 12 mothers and 5 community health workers with experience of the intervention, and 2 members of the implementing research team who supervised the implementation of the intervention in refugee camps.

### 2.3. Intervention

The intervention primarily comprised of pictorial tool-assisted counselling on early child development and maternal mental health. Core content of the intervention for children focused on, mental stimulation and responsive caregiving, and nutritional messages for improving cognitive and physical health of children. Core content of the intervention for mothers focused on behavioural activation strategies to prevent depression by improving self-care, social support and daily functioning.

The ECD intervention comprised of four counselling sessions, delivered on quarterly basis by community health workers. The counselling content for each session incorporated age-appropriate mental stimulation activities and nutritional messages for promoting child development. The intervention sessions were delivered by the Community Health Workers (CHWs) at the place of residence of mothers. The counselling material was adapted to the culture of forcibly displaced Myanmar nationals.

Community health workers, having at least 10 years of formal education, were recruited from the refugee population for intervention delivery to minimize language barrier and other challenges.

CHWs were given 4 -day training on the intervention content and delivery using the flipbooks. Complete details of the intervention are provided in the study protocol(11).

### 2.4. Implementation framework

Proctor and colleagues’(13) implementation framework was used to explore the acceptability, adoption, appropriateness, feasibility, and cost in the intervention arm. Acceptability was defined as the satisfaction of the CHWs, mothers and their families with the intervention. Adoption was defined as the uptake or implementation of the intervention strategies by mothers in their daily routine. Appropriateness was defined as the perceived fit, relevance, compatibility of the ECD care package, content and mode of delivery for the mothers and CHWs. Feasibility was defined as the ease of implementation of the intervention for the CHWs and mothers and the challenges experienced by both regarding the implementation or uptake of the intervention. Cost was defined as the expenses incurred on implementation of the care package in both arms from a healthcare provider perspective(13).

### 2.5. Data Collection and analyses Quantitative data

Quantitative data was extracted primarily from the project records on multiple selected indicators for CHWs and mother-child dyads. For the CHWs, quantitative data on number and proportion of CHWs recruited for intervention delivery and feasibility of training was recorded. For the mother-child dyads, quantitative data on number and proportion of participant recruitment rate, reasons for refusals, adherence to intervention strategies, follow-up attendance rate, and referral rate. Data was analysed using descriptive analyses in SPSS version 26.

#### Qualitative data

Semi-structured interviews were conducted with mothers in household settings by female researchers. A topic guide was developed in consultation with local stakeholders in Burmese language. Interviews were conducted after completion of endpoint assessments. Each interview lasted for 30-45 minutes on average. Data was collected on facilitators, barriers and recommendations to improve implementation of the ECD care package in refugee camps of Bangladesh. Interviews were transcribed in English language. Data was analysed using thematic analyses(14) by two researchers, using deductive analyses whereby emergent codes from data were added to pre-defined thematic codebook.

#### Incremental cost-effectiveness data collection and analyses

Costs were extracted from budget and expense reports of the project, using the healthcare provider perspective on implementation of the ECD care package and using the ingredients approach(15). The health services costs comprised of both the capital and recurrent costs incurred in intervention implementation and costs incurred to standardize control arm. Capital costs were calculated using the costs of training CHWs in both arms, developing contextualized training manuals, flipbook, for intervention arm, and costs incurred on anthropometric scales in both arms. Recurrent costs were calculated by estimating the costs for brochures in intervention arm, communication equipment, recording tools and conveyance incentives provided to CHWs in both arms. No discounting was required as the time horizon was less than one year. The BDT to USD currency exchange rate given is for the year 2021.

The primary treatment outcome of the trial was the reduction in proportion of children with two or more delays in the five domains i.e., communication, gross motor, fine motor, problem solving and personal social at endpoint of 12 months of child’s age, measured using Ages and Stages Questionnaire-3 (ASQ-3). Data on effectiveness outcome was extracted from the trial data and is described in the trial outcome paper (publication in process). Incremental cost-effectiveness ratio was calculated to estimate the incremental cost-effectiveness of the ECD care package in reducing the proportion of children with two or more developmental delays by one percentage point..

### 2.6. Ethical considerations

Ethical approval for the study was obtained from (1) Bangladesh Medical Research Council for research under reference number BMRC/NREC/2016-2019/843 and (2) Refugee, Rehabilitation and Repatriation Commission for project implementation under reference number ShoTraProKa/RHU/ARK Foundation/13/2019/589. Written informed consent was obtained from mothers for their participation in the study. Confidentiality of the participants was ensured in compliance with Good Clinical Practices.

### 2.7. Data management

Quantitative data was collected in hard copy by the CHWs and was entered by the data entry team in SPSS. All identifying information of participants was removed and stored separately by the data entry team. Descriptive analyses were used to ensure the reliability and integrity of data. The dataset was stored in password protected computers. The research team had access to the anonymized data.

All identifiable information was removed from the transcriptions and each participant was assigned an ID to ensure confidentiality and privacy of participants. Anonymised data was stored in paper format in locked filing cabinets in the field office.

## 3. Results

### 3.1. Acceptability

22 CHWs out of 66 potentially eligible candidates were recruited to deliver the intervention to mother-child dyads in the refugee camps. All selected CHWs agreed to deliver the intervention and found the intervention delivery method to be acceptable.

*“When i was given the opportunity to contribute to improvement of children’s and mothers’ health in our community, I thought it would be a very useful way to contribute towards rebuilding our community (CHW-age 26-30 years)*

Of the eligible 355 mothers, majority of mothers found the program acceptable and agreed to participate. Only 2% of potentially eligible mothers refused to participate in the study due to multiple reasons.

*Most of the mothers agreed to participate in the study. Very few mothers refused because they either intended to move from the area or were uncertain about their future plans. (CHW-age 21-25 years)*

The CHWs reported some cultural barriers that hindered the anthropometric measurements of children.

*“Some mothers didn’t feel comfortable letting us measure their child due to cultural myths that measuring a child’s height and weight impedes his growth but they agreed when we explained the importance of monitoring child’s height and weight” (CHW-age 26-30 years)*

Most of the mothers were very forthcoming about their mental health difficulties and found the assessment sessions to be very cathartic experience.

*“They asked me some questions about how I felt and It was nice that someone was asking me about my wellbeing as it’s a very tough situation we are in and it felt good to talk to someone. I could talk to them easily because they were female from our own community and we knew them and they asked me these questions in a very kind manner. When I didn’t understand a message, they explained it to me” (Mother, age 26-30 years)*

Given the fact that the counselling sessions were delivered to mothers at their residing places, the mothers reported that their family members were very keen on participation of the mother-child dyads in the sessions. *My mother-in-law and husband were also very happy that someone was coming to our home to give us information that could improve my health and health of my child. We thought that the use of picture book (pictorial flipbook) was a very interesting way to tell us about different ways in which we can improve our child’s health. (Mother, age 21-25 years)*.

### 3.2. Feasibility

Implementation team reported the engagement of camp administration to be a facilitating factor in intervention implementation. *“The engagement of camp administration was especially helpful in implementation, as people were more comfortable in interacting with us as they knew we were affiliated with a credible and recognized authority in the area.” (Project coordinator)*.

CHWs identified the potentially eligible mother-child dyads for participation in the study using birth records maintained by the health department. Around 360 mother-child dyads were approached by the CHWs and 355(99%) were identified to be eligible.

*“The participants were recruited using door-to-door recruitment approach in community settings, which is why the recruitment took around 3 months. It was a bit challenging but it helped us achieve our recruitment milestones efficiently” (Monitoring officer)*

The counselling sessions were delivered at residing places of mothers, which was reported to be a facilitator for both the mothers and the CHWs.

*“We were able to effectively deliver sessions because we coordinated with the mothers to deliver sessions at a time that was convenient for them. It was workable for us because we lived in the same camp as our assigned mother-child dyads for intervention delivery so we could arrange our intervention delivery schedule to suit the preferences of mothers. (CHW, age 26-30 years)*.

CHWs reported the carrying of heavy anthropometric scales to be a challenge for them. *“These scales are heavy and we had to carry these around to each household on every visit. So sometimes I enlisted the help of my young nephew to help me carry this stuff. (CHW, age 26-30 years)*

The CHWs counselled around 2 mother-child dyads per day on average, as they resided in the same camps as that of intervention participants assigned to them. Counselling sessions were delivered with the help of pictorial tool and each session was 30 minutes long on average.

*“The counselling sessions were delivered using the pictorial flipbook, which the mothers found very easy and interesting. It was easier for them to understand the messages with the help of images in the pictorial book and they were able to relate with the pictures, which is why sessions were completed on time otherwise it may have taken longer to get them to understand the concepts. (CHW-aged 31-35 years)*

*“The messages were very simple and easy to understand. I had to look at the picture in the book and I quickly understood how to implement a physical activity with my child or how to take care of myself easily”. (Mother, age 26-30 years)*

### 3.3. Appropriateness

The engagement of stakeholders helped ensure the appropriateness of the intervention for delivery agents and target population.

*“We engaged our stakeholders throughout the intervention adaptation procedure and it helped ensure that the intervention was culturally appropriate and was relevant to the target population” (Project coordinator)*

The CHWs and the mothers reported the delivery of intervention by refugee women (CHWs) from the same population as that of the mother-child dyads to be an appropriate mechanism of intervention delivery.

*“All the CHWs who were selected had one thing in common; they wanted to improve the wellbeing of the vulnerable mothers and children in the refugee camps, so I think the recruitment procedure was effective in selecting the suitable people for implementing the intervention in community”. (CHW-age 26-30 years)*.

*The community health workers were from our own community, we knew them and they knew us and the challenges we face in our daily lives so when they approached us about the study, we felt very comfortable participating in the program as we knew our CHW was genuinely interested in improving the health of our child. I decided to take part in the program and completed it due to our CHW (Mother-age 31-35 years)*

### 3.4. Adoption

The mother-child dyads were followed-up on quarterly basis for child’s anthropometric measurements and the delivery of sessions. Record of the follow-up sessions show that around 341 (98%) mothers were able to attend all four sessions. The high attendance rate is attributed to the delivery of counselling sessions at residing places of mother-child dyads.

*“The CHW was very flexible in scheduling the session as per my convenience. She understood that I am a new mother with a lot of responsibilities so she delivered the sessions at a time when I was free and she always came to our house to deliver the session so I attended all the sessions. (Mother-age 31-35 years)*

Mothers and children were referred to a public doctor if the CHW felt they needed specialist attention. Around 4(1%) mother-child dyads were referred to the clinic. However, most of the mothers 3 (75%) refused the referral due to barriers involving inability to visit a health facility due to transportation challenges, cost of consultation and restrictions imposed on mobility as well as services that could be availed by them outside their residing camps.

*“Our mobility is restricted within camps; we cannot move around easily and we don’t have the money to pay for the fare to a clinic and then to pay the doctor to see our child. (Mother- age 31-35 years)*

Some of the reported barriers in intervention implementation were other work commitments of mothers such as domestic work and poor wellbeing of mother herself.

*“We talked to the CHWs working in intervention arm and they reported that some mothers wanted medicines or vitamin supplements. But it was only in very few cases and it’s understandable that some people would want medicines as they are living in very tough circumstances but most of the mothers didn’t expect this as they knew the CHWs were from their own community and were only trained in delivering the intervention and didn’t have access to this stuff. (Monitoring officer)*

The intervention strategies were found to be very engaging and feasible to implement with children. Most of the mothers (i.e., 98%) reported to have done all the activities suggested in the intervention.

*“The sessions were short which was good because we have a lot of things to do to manage livelihood in our camps. It was easy to do the activities they taught us in daily routine, these were very simple and easy and did not require any extra equipment. It also made me spend some dedicated time with my child” (Mother- age 26-30 years)*

*They told me to do a bunch of activities in my daily routine for improving my wellbeing, such as talking to a close friend or family member. These seemed very simple but made me feel better. I actively tried to implement these in routine (Mother- age 26-30 years)*

### 3.5. Cost and Cost-effectiveness Healthcare provider costs

Table 1 summarizes the capital and recurrent unit costs incurred on implementation of the ECD care package from a healthcare provider perspective and the costs incurred on standardizing the control arm. The total capital costs incurred in intervention and control arms were 725 USD and 422 USD respectively, while the recurrent costs were 1716 USD and 1009.5 USD. The total costs incurred in intervention arm were higher at 2441 USD as compared to the total costs incurred in the control arm at 1431 USD. Costs associated with research relevant procedures and supervision were not added to the costs of intervention implementation. The cost of intervention implementation per child was 7 USD.

**Table 1.**
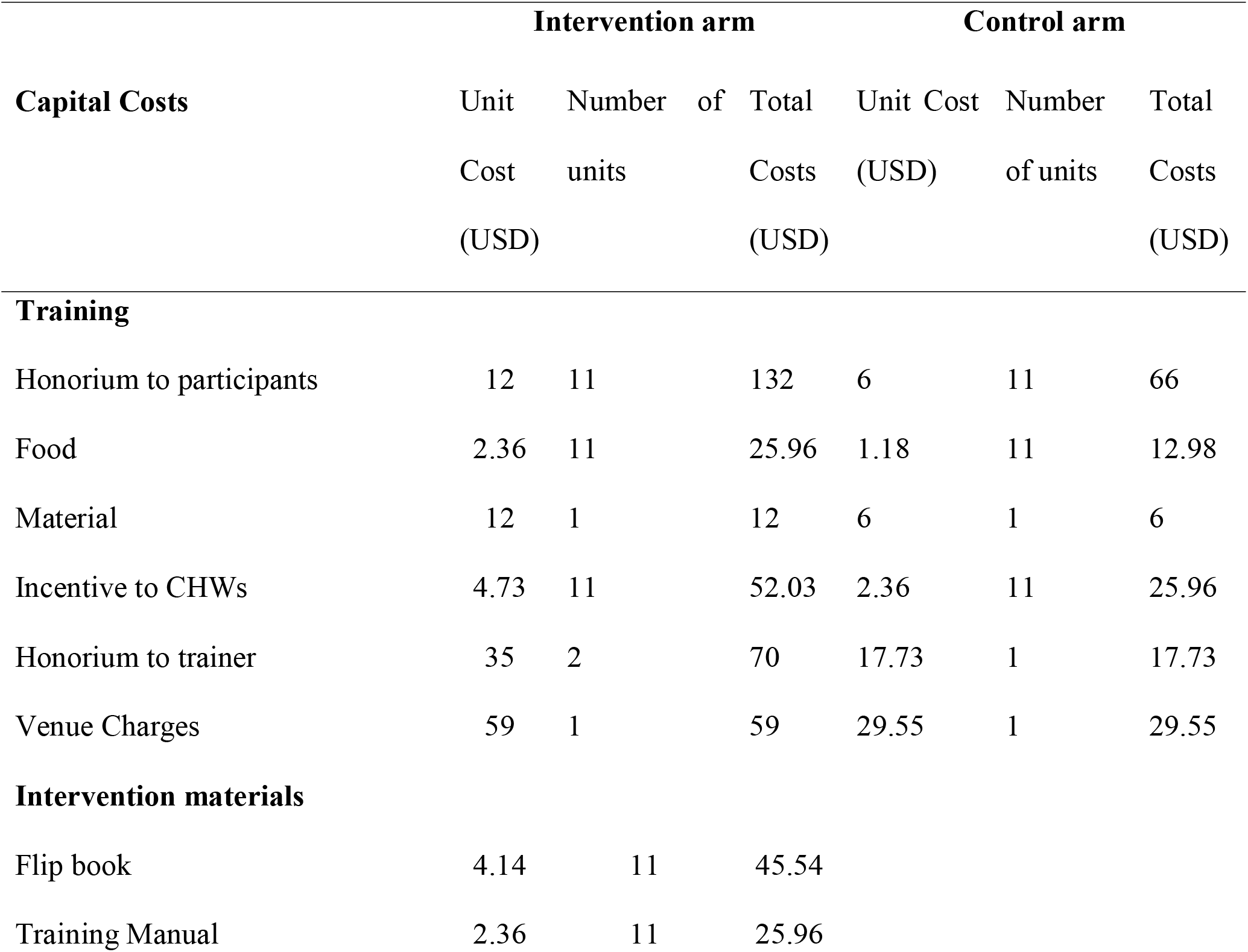
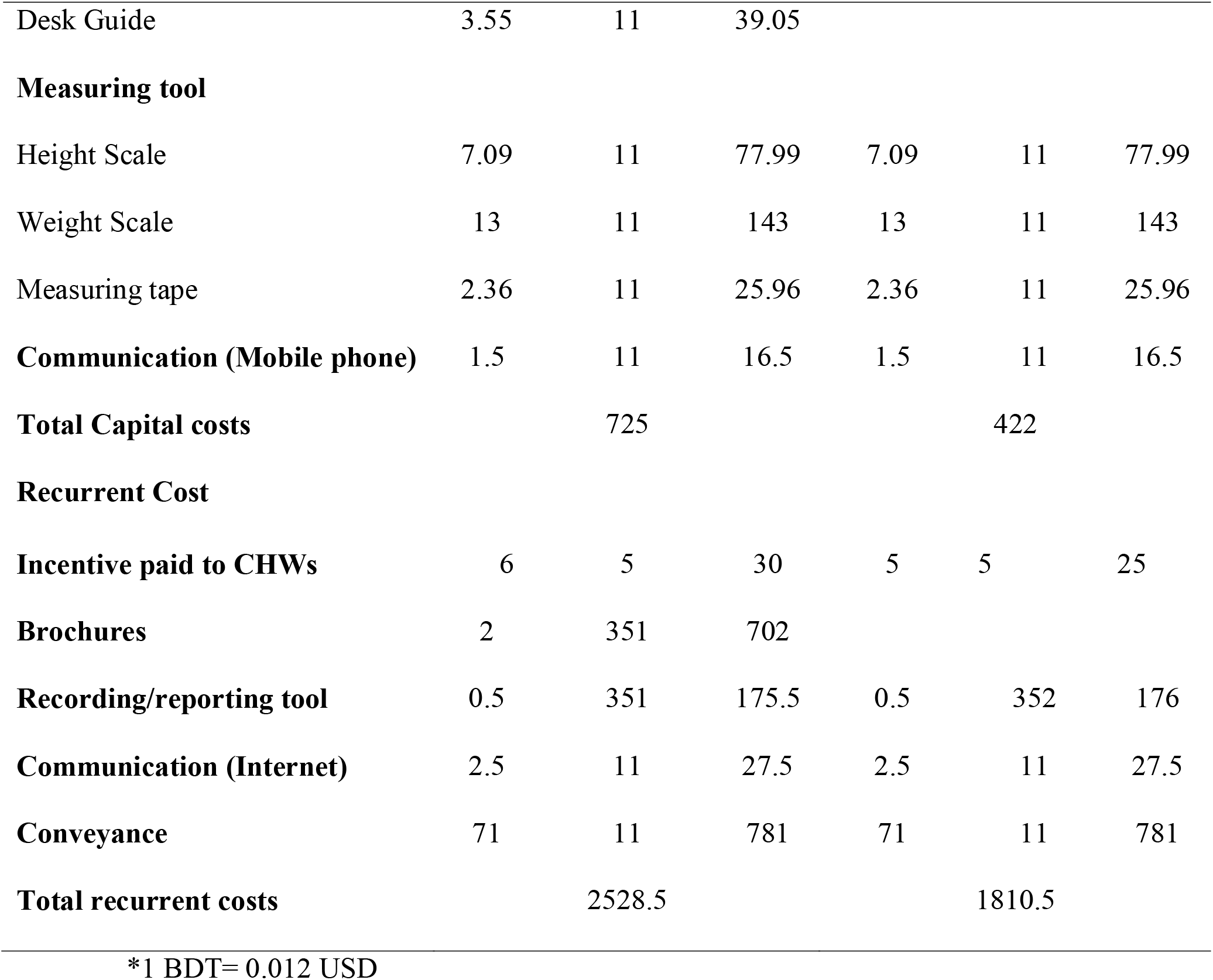
Costs of implementing ECD care package in intervention and costs of standardizing control arm.

#### Effectiveness outcomes

The ECD care package was effective in reducing proportion of children with two or more developmental delays in intervention arm as compared to control arm at endpoint of 12 months of child’s age. The crude difference in the proportion of two or more developmental delays reduced across arms was 16% at endpoint (described in the trial outcome paper – publication in process)

#### Incremental cost-effectiveness analysis

The Incremental Cost Effectiveness Ratio (ICER) provided an estimate for the incremental cost incurred in reducing the proportion of children with two or more developmental delays by one percentage pointat endpoint. ICER for reduction in proportion of children with two or more developmental delays was calculated using the difference in costs across two arms divided by the difference in effectiveness outcomes across both arms (2441-1431/16) which yielded an incremental cost of 63 USD in reducing the proportion of children with two or more developmental delays by one percentage point

## 4. Discussion

The process evaluation proved to be crucial in gaining insight into the experiences of mothers, CHWs and research team, and facilitators and barriers experienced in implementation of the early child development and maternal depression intervention for forcibly displaced Myanmar nationals in Bangladesh. Despite the challenges of implementing a health initiative in a refugee setting devoid of basic infrastructure and resources, the intervention was indicated to be acceptable, feasible, appropriate, adoptable and cost-effective.

The recruitment of community health workers from within the community of the target population helped address the language barriers and develop a trusting bond between the delivery agents and the participants. The difficult circumstances within the refugee camps are generally a constraint for any outsider to work effectively. Recent studies conducted with refugee population in Bangladesh reported a high turnover rate of staff members recruited to deliver interventions in the refugee camps due to security concerns and language barriers of staff members delivering the intervention, however these challenges were pre-empted in this study by recruiting community health workers from within the refugee population which facilitated the rapport building and successful intervention delivery(6, 16). The engagement of the key public health officials was helpful in accessing refugee camps and delivering the ECD and mental health care within the refugee camp context. Other documented experiences from Bangladesh have reported similar positive effect of engaging key local personnel for smooth implementation of health initiatives and ECD programs(17, 18).

Contextualizing the content and delivery of intervention through, a multi-stakeholder working group, was a key facilitator. Similar beneficial effects of contextualizing the ECD interventions have been reported by other studies conducted in Bangladesh (6, 16). The feasibility of the intervention delivery and adoption could be attributed to the use of brief, simple and relatable pictorial messages used to deliver intervention to mothers, the short duration of intervention sessions which helped mothers manage time for the intervention along with their other responsibilities, and the engagement of the family members along with the mothers by the CHWs. In various other studies; adaptation of the intervention to make it brief and appropriate for delivery in short duration has been suggested as a key recommendation for successful implementation of ECD programs. (9, 19, 20).

Keeping the intervention activities simple with minimal requirements of education, skills, material resources, has been found to be crucial in successful adoption of the recommended activities in daily routine. Other studies conducted in Bangladesh have also reported that merging the intervention curriculum into daily routine of mother-child dyads can prove to be crucial for the successful adoption of program activities (17).

The dose received of the intervention being delivered in home-based setting (i.e., within the residing refugee camp of the mother) was exceptionally high as compared to when this intervention was evaluated in facility-based setting in Pakistan, where only 34% of mothers completed all three follow-up visits to the facility(21). Previous studies conducted in Bangladesh to improve early child development also report that intervention delivery in households of participants and ensuring engagement of extended family members, especially the in-laws, are crucial facilitating factors for mothers(17). As the intervention was delivered by women in the refugee camps and was not health facility based, therefore despite Covid-19 challenges, the implementation was not much hindered.

Cultural myths about children’s anthropometric measurements were a key challenge reported by CHWs in intervention implementation. Similar findings have been reported by other studies conducted in Bangladesh, whereby supernatural myths inconsistent with the intervention messages were a barrier in the delivery(17).

In this study, the cost incurred in reducing the proportion of children with two or more developmental delays by one percentage point was 63 USD which compares favourably with findings reported in other studies conducted in non-refugee settings of middle- and high-income countries. A study conducted in primary healthcare settings of China to evaluate the effectiveness and cost-effectiveness of an intervention to improve child development found the implementation cost of intervention per child to be $51(22). A study conducted in Pakistan that integrated delivery of ECD care within a maternal and neonatal program(10), found the implementation cost of intervention per child to be 4 USD per month, which is comparable with the per child implementation cost of 7 USD found in our study. Similarly, parenting programs implemented in Ireland and the United Kingdom found the ICER to change one point score in Eyeberg intensity score ranged from 87 USD(23) to 116 USD(24) respectively. To the best of our knowledge cost-effectiveness analyses of an ECD package for reducing proportion of developmental delays among children has not been evaluated. The cost can be further reduced by achieving economies of scale if this intervention is scaled to a larger population. Furthermore, the current cost-effectiveness analysis does not account for proportion of children with one developmental delay prevented which could further decrease the incremental cost-effectiveness of intervention in reducing developmental delays. A health intervention that costs less than the per-capita gross domestic product (GDP) of a given country is considered to be very cost-effective (22, 25). The ICERs of our intervention were below the per-capita GDP of Bangladesh ($1855 in 2019), which implies that the ECD care package is scalable.

A limitation of the study was the inability to establish more effective referral links with the general health care in the district. Mothers reported an inability to visit any health facility outside refugee camps because of direct, indirect costs associated with the visit, restricted mobility, and access to health services. Other studies conducted in refugee settings have also reported this challenge, indicating the need to incorporate a more effective method to establish referral link with the health sector to ensure adequate care is provided to mother-child dyads needing specialist attention(6). Furthermore, the cost-effectiveness analysis was only conducted from the healthcare-provider perspective, which might have led to the intervention being more cost-effective as the costs incurred by participants were not accounted for as these were assumed to be negligible for home-based delivery.

## 5. Conclusion and implications for future

This study provides evidence for the cost-effectiveness of ECD care package in refugee settlements and the resource allocation required for ensuring sustainable delivery of the intervention in refugee camps. The community-based delivery of care through trained refugee women has the potential to reduce excessive dependence on sustained external inputs for continuing the care. The enabled refugee women will remain available in their respective communities as a continued source of advice. This ECD care package can be incorporated in the refugee health services in the refugee camps. Future studies can implement and evaluate the effectiveness of this ECD care package when implemented at scale in other refugee camps.

## Data Availability

All data produced in the present study are available upon reasonable request to the authors

## Acknowledgments

We acknowledge the Directorate General of Health Services and Former Director of Primary Health Care Support, Ministry of Health and Family Welfare, for their technical guidance. We are also grateful to the Refugee, Rehabilitation and Repatriation Commission at Cox’s Bazar for their support and approval. Heartiest thanks are extended to the Civil Surgeon of Cox’s Bazar and the Upazila Health and Family Planning Officer of Ukhiya for their kind support with the project.

## References

1. Migration IOf. Rohingya humanitarian crisis response—External update, 4–10 May 2018 2018 [Available from: (https://reliefweb.int/report/bangladesh/iom bangladesh rohingya humanitarian crisis respo nse external update 4 10 may 2018.

2. Refugees UNHCf. [Available from: http://www.unhcr.org/globaltrends2016/

3. Walker SP, Wachs TD, Grantham-McGregor S, Black MM, Nelson CA, Huffman SL, et al. Inequality in early childhood: risk and protective factors for early child development. The lancet. 2011;378(9799):1325–38.

4. Grantham-McGregor S, Cheung YB, Cueto S, Glewwe P, Richter L, Strupp B, et al. Developmental potential in the first 5 years for children in developing countries. The lancet. 2007;369(9555):60–70.

5. Patel V, Rahman A, Jacob K, Hughes M. Effect of maternal mental health on infant growth in low income countries: new evidence from South Asia. Bmj. 2004;328(7443):820–3.

6. Sarker M, Saha A, Matin M, Mehjabeen S, Tamim MA, Sharkey AB, et al. Effective maternal, newborn and child health programming among Rohingya refugees in Cox’s Bazar, Bangladesh: Implementation challenges and potential solutions. PloS one. 2020;15(3):e0230732.

7. Ahmed S, Lee S, Shommu N, Rumana N, Turin T. Experiences of communication barriers between physicians and immigrant patients: A systematic review and thematic synthesis. Patient Experience Journal. 2017;4(1):122–40.

8. Khan MA, Owais SS, Maqbool S, Ishaq S, Khan HJ, Minhas FA, et al. Is integrated private-clinic based early child development care effective? A clustered randomised trial in Pakistan. BJGP open. 2018;2(2).

9. Hamadani JD, Huda SN, Khatun F, Grantham-McGregor SM. Psychosocial stimulation improves the development of undernourished children in rural Bangladesh. The Journal of nutrition. 2006;136(10):2645–52.

10. Yousafzai AK, Obradović J, Rasheed MA, Rizvi A, Portilla XA, Tirado-Strayer N, et al. Effects of responsive stimulation and nutrition interventions on children’s development and growth at age 4 years in a disadvantaged population in Pakistan: a longitudinal follow-up of a cluster-randomised factorial effectiveness trial. The Lancet Global Health. 2016;4(8):e548–e58.

11. Al Azdi Z IK, Khan MA, Khan N, Ejaz A, Khan MA, Warraitch A, Jahan I, Huque R. Effectiveness of an integrated care package for Refugee mothers and children: protocol for a cluster randomized controlled trial. JMIR Research Protocols. 2021.

12. Moore GF, Audrey S, Barker M, Bond L, Bonell C, Hardeman W, et al. Process evaluation of complex interventions: Medical Research Council guidance. bmj. 2015;350.

13. Proctor E, Silmere H, Raghavan R, Hovmand P, Aarons G, Bunger A, et al. Outcomes for implementation research: conceptual distinctions, measurement challenges, and research agenda. Administration and Policy in Mental Health and Mental Health Services Research. 2011;38(2):65–76.

14. Gavin H. Thematic analysis. Understanding research methods and statistics in psychology. 2008:273–82.

15. Levin HM, McEwan PJ. Cost-effectiveness analysis as an evaluation tool. International handbook of educational evaluation: Springer; 2003. p. 125–52.

16. Mahmuda M, Miah MAA, Elshazly M, Khan S, Tay AK, Ventevogel P. Contextual adaptation and piloting of Group Integrative Adapt Therapy (IAT-G) amongst Rohingya refugees living in Bangladesh. Intervention. 2019;17(2):149.

17. Hamadani JD, Nahar B, Huda SN, Tofail F. Integrating early child development programs into health and nutrition services in Bangladesh: benefits and challenges. Annals of the New York Academy of Sciences. 2014;1308(1):192–203.

18. Ahmed R, Aktar B, Farnaz N, Ray P, Awal A, Hassan R, et al. Challenges and strategies in conducting sexual and reproductive health research among Rohingya refugees in Cox’s Bazar, Bangladesh. Conflict and Health. 2020;14(1):1–8.

19. Tofail F, Hamadani JD, Mehrin F, Ridout DA, Huda SN, Grantham-McGregor SM. Psychosocial stimulation benefits development in nonanemic children but not in anemic, iron-deficient children. The Journal of nutrition. 2013;143(6):885–93.

20. Nahar B, Hamadani JD, Ahmed T, Tofail F, Rahman A, Huda S, et al. Effects of psychosocial stimulation on growth and development of severely malnourished children in a nutrition unit in Bangladesh. European Journal of Clinical Nutrition. 2009;63(6):725–31.

21. Khan MA, Owais SS, Ishaq S, Walley J, Khan HJ, Blacklock C, et al. Process evaluation of integrated early child development care at private clinics in poor urban Pakistan: a mixed methods study. BJGP open. 2017;1(3).

22. Shi H, Li X, Fang H, Zhang J, Wang X. The Effectiveness and Cost-effectiveness of a Parenting Intervention Integrated with Primary Health Care on Early Childhood Development: a Cluster-Randomized Controlled Trial. Prevention Science. 2020;21:661–71.

23. O’Neill D, McGilloway S, Donnelly M, Bywater T, Kelly P. A cost-effectiveness analysis of the Incredible Years parenting programme in reducing childhood health inequalities. The European Journal of Health Economics. 2013;14(1):85–94.

24. Edwards RT, Céilleachair A, Bywater T, Hughes DA, Hutchings J. Parenting programme for parents of children at risk of developing conduct disorder: cost effectiveness analysis. Bmj. 2007;334(7595):682.

25. Hutubessy R, Chisholm D, Edejer TT-T. Generalized cost-effectiveness analysis for national-level priority-setting in the health sector. Cost effectiveness and resource allocation. 2003;1(1):1–13.

